# Symbolic transfer entropy reveals the age structure of pandemic influenza transmission from high-volume influenza-like illness data

**DOI:** 10.1101/19005710

**Authors:** Stephen M Kissler, Cécile Viboud, Bryan T Grenfell, Julia R Gog

## Abstract

Existing methods to infer the relative roles of age groups in epidemic transmission can normally only accommodate a few age classes, and/or require data that are highly specific for the disease being studied. Here, symbolic transfer entropy (STE), a measure developed to identify asymmetric transfer of information between stochastic processes, is presented as a way to determine which age groups drive an epidemic. STE provides a ranking of which age groups dominate transmission, rather than a reconstruction of the explicit between-age-group transmission matrix. Using simulations, we establish that STE can identify which age groups dominate transmission, even when there are differences in reporting rates between age groups and even if the data is noisy. Then, the pairwise STE is calculated between time series of influenza-like illness for 12 age groups in 884 US cities during the autumn of 2009. Elevated STE from 5-19 year-olds indicates that school-aged children were the most important transmitters of infection during the autumn wave of the 2009 pandemic in the US. The results may be partially confounded by higher rates of physician-seeking behaviour in children compared to adults, but it is unlikely that differences in reporting rates can explain the observed differences in STE.

## 1. Introduction

Age is a key predictor of a person’s rate of both acquiring [1, 2, 3, 4, 5, 6] and transmitting [7, 8, 9] influenza. Children tend to contribute more to influenza transmission than adults do [4, 7, 8], but the precise epidemiological roles of different age groups can shift from season to season [10] and may change markedly in pandemic years [11]. From a public health perspective, untangling the relative roles of different age groups could help guide targeted vaccination strategies [7, 12, 13, 14] and other age-related interventions, like the selective closure of schools [15, 16, 17]. However, data with sufficient resolution to identify detailed epidemiological relationships between age groups has so far been scarce, and even when such data exist, current methods are insufficient for reliably uncovering those relationships.

Electronic medical records (EMRs) help address the issue of data scarcity by providing high-volume influenza-like illness (ILI) incidence data with detailed age structure [18]. EMRs are routinely produced by physicians for insurance purposes during the majority of outpatient visits in the United States [18]. Since EMRs generally contain syndromic illness classifications, EMR-based estimates of influenza incidence are subject to noise from non-ILI respiratory infection. EMR-based disease incidence estimates are also subject to geographic and demographic variation in physician-seeking behaviour. Laboratory-confirmed influenza cases, as collected routinely by the Centers for Disease Control and Prevention (CDC) [19], provide more specific estimates of influenza incidence, but at substantially lower volume. Influenza incidence estimates from online search platforms and social media websites like Google [20] and Twitter [21] can provide massive amounts of data, but these sources’ reliability has been called into question, and they lack detailed age information [22]. Dedicated online platforms such as FluNearYou in the US and FluSurvey in the UK, which gather reports of ILI symptoms from community volunteers [23, 24], hold some promise for supplementing traditional ILI data streams [25, 26, 27], but represent a relatively small convenience sample of the population. So, while other data sources exist, EMRs offer a relatively promising and so-far underutilised source of fine-scale data on influenza incidence in the United States [18, 22].

Previous attempts to infer the relative importance of different age groups for the transmission of influenza have sought to either reconstruct the explicit next-generation matrix (NGM) [3, 28, 29] or to infer the relative risk of infection between age groups [4]. The NGM-based methods have only been applied to scenarios with at most two age groups (children and adults), in part because they require strong assumptions about the structure of the next-generation matrix which become increasingly unrealistic as the number of age classes grows. The relative risk method [4] has been used to rank the importance of five age groups for the transmission of influenza, but requires data with high specificity for influenza, effectively precluding ILI datastreams and the use of EMRs in particular.

Symbolic transfer entropy (STE) [30] offers a way to infer the relative transmissive importance of possibly many age groups from ILI data. STE is an extension of transfer entropy (TE) [31], which measures the amount of information the past states of one stochastic process provide about the transition probabilities of another. Intuitively, the TE is a measure of the amount of information “transferred” from one stochastic process to another. To compute the STE, a time series is symbolised using a scheme that encodes its qualitative structure in a low-dimensional space, and then the TE is calculated from the relative frequencies of these symbols. The symbolisation scheme makes the STE robust to minor point-wise noise and to systematic shifts in amplitude, which in the context of EMR ILI data might arise from the presence of non-influenza ILI cases and from differences in reporting rate between age groups. These benefits come with the trade-off of requiring relatively large amounts of data compared to existing methods for inferring the age structure of disease transmission. STE has been used to study epileptogenic neural signals and the dissemination of information through social networks [30, 32], but to our knowledge has not been systematically evaluated as a means of providing insight into infectious disease transmission. TE and STE are similar to other model-free methods that measure shared information and so-called ‘causal’ relationships between stochastic processes, including mutual information [31], Granger causality [33], and convergent cross mapping [34]. Permutation entropy, a related measure, has recently been used to quantify the predictability of infectious disease outbreaks [35].

Here, we use influenza-like outbreak simulations to demonstrate that STE reliably identifies the age groups that drive influenza transmission. Then, we utilise an EMR-based dataset capturing ILI incidence from 884 ZIP (postal) codes and 12 age classes across the United States to rank the relative importance of the various age groups in the transmission of the autumn wave of the 2009 A/H1N1pdm influenza pandemic in that country. We conclude that school-aged children (5–19 year-olds) were disproportionately responsible for transmitting influenza to infants through working-age adults in the autumn of 2009, in broad agreement with other findings. Our work demonstrates that STE could serve as an important tool for the detailed epidemiological analysis of age structure, especially as EMR data become more prevalent.

## 2. Materials and Methods

### 2.1. Data

The data come from a convenience sample of CMS-1500 electronic medical claims forms submitted by primary care physicians across the US and maintained by SDI health (now IQVIA). Each claim is associated with a single outpatient visit, and includes one or more ICD-9 codes [36] listed by the physician that describe the patient’s illness. The overall sample is thought to capture over 50% of all outpatient visits in the US in 2009 [18]. The records are binned weekly and aggregated geographically by the first three digits of the ZIP (postal) code of the practice from which they are submitted [37]. These three-digit ZIP codes will be referred to simply as ‘ZIPs’ (not to be confused with the finer five- or ten-digit ZIP codes, also assigned to many mailing addresses in the US [36]). Time series of weekly influenza-like illness (ILI) incidence are created by extracting claims with a direct mention of influenza, or fever combined with a respiratory symptom, or febrile viral illness (ICD-9 487-488 OR [780.6 and (462 or 786.2)] OR 079.99), following Viboud *et al*. (2014) [18]. For each ZIP, the number of ILI cases in each week is divided by the total number of patients who visited a physician in that ZIP during that week, yielding an ‘ILI ratio’ time series. There are 884 ILI ratio time series, one for each ZIP in the lower 48 US states, each spanning 52 weeks from the week commencing 4 Jan 2009 through the week commencing 27 Dec 2009. The correspondence between the SDI-ILI dataset and reference influenza surveillance data from the US Centers for Disease Control and Prevention (CDC) is described in depth by Viboud *et al*. (2014) [18].

### 2.2. Symbolic transfer entropy

If *I* and *J* are discrete-state and discrete-time random processes such that *i*_*t*_ and *j*_*t*_ are the states of processes *I* and *J* at time *t*, then the transfer entropy (TE) from process *J* to process *I* is defined as

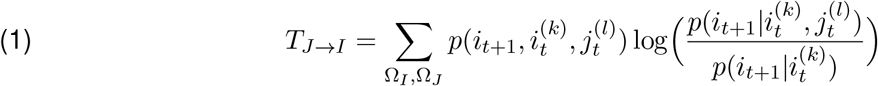

where 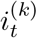 is shorthand notation for the *k*-step history of process *i*, (*i*_*t*_, …, *i*_*t*−*k*+1_), and similarly 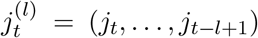. The logarithm has base 2, so that the transfer entropy is measured in bits. The sum is over all possible combinations of states 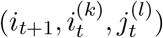, where 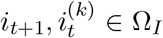 and 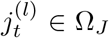, and Ω_*I*_ and Ω_*j*_ are the state spaces for processes *I* and *J*. Eq. 1 is a Kullback-Leibler divergence that measures how much process *I* deviates from the generalised Markov property 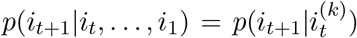, given the last *l* states of process *J*. In practice, the histories are often fixed at length 1 (*k* = *l* = 1) and the probabilities are estimated from simple counts of the observed data [31].

The TE is limited in that it is only defined for stochastic processes with a discrete state space. Staniek and Lehnertz (2008) [30] introduce symbolic transfer entropy (STE) as a way to calculate information transfer between time series processes that have continuous- or near-continuous state spaces. Motivated by the insight that the relative amplitudes of subsequent observations from these sorts of processes may provide enough information to reveal interactions between them, they propose symbolising the time series based on ordered *m*-tuples of observations (Fig. S1). This reduces the (near-)continuous state space of the original stochastic process to a discrete set of *m*! symbols. In practice, *m* is often chosen to be 2 or 3, giving a state space of 2 or 6 symbols, respectively. For *m* = 3, we also tested the effect of collapsing the two concave-up and the two concave-down symbols into a single symbol each, resulting in a smaller state space (four *vs*. six symbols) while capturing a similar level of qualitative detail. Details on the symbolisation of time series and the empirical calculation of the STE are provided in the Supplemental Information.

### 2.3. SIR epidemic simulation model

For simulations with just two age classes, we use a stochastic SIR model implemented using the Gillespie algorithm [38]. For all simulations, the basic reproduction number *R*_0_ is set at 1.5, consistent with estimates of the basic reproduction number of 2009 A/H1N1 pandemic influenza [39, 40]. We consider a population size of *N* = 1, 000 split evenly between classes 1 and 2, so that *N*_1_ = *N*_2_ = 500 (age groups with different population sizes are also considered in the Supplemental Information). The expected time to recovery 1*/γ* is assumed constant for all age groups and is set at 7 days, which is consistent with estimates of the infectious period for 2009 pandemic influenza [40]. Table S1 gives the rates at which individuals of each class stochastically progress from susceptible to infected to recovered. Infections are binned into week-long intervals, and Poisson noise is added to simulate non-influenza influenza-like illness. Fig. S7 depicts five incidence time series produced using the model. Full details on the model and simulation procedure are given in the Supplemental Information.

### 2.4. Poisson epidemic simulation model

For more than two age classes, the full stochastic SIR model becomes too computationally demanding for repeated simulations to be practical. So, we also define an outbreak simulation model based on a self-exciting Poisson process, similar to [41]. We choose the time units *t* to match the mean generation interval of the infection, which we set at 3.5 days [42]. To generate epidemics, we use a stepwise-constant effective reproduction number *R*_*t*_, such that *R*_*t*_ = 1.5 for the first four weeks (eight generations) of the outbreak and *R*_*t*_ = 0.8 thereafter. Infections are binned from the half-week generations into week-long intervals, and additional Poisson noise is added to each bin to simulate non-influenza influenza-like illness. For simulations with two age classes, the Poisson model yields epidemics of similar length and magnitude as the two-age-class SIR model (compare Figs S7 and S8), and yields comparable STE inferences (see Fig. 1), which suggests that the Poisson model is an acceptable approximation to the stochastic SIR model. Full details on the implementation of the Poisson model are given in the Supplemental Information.

**Figure 1.**
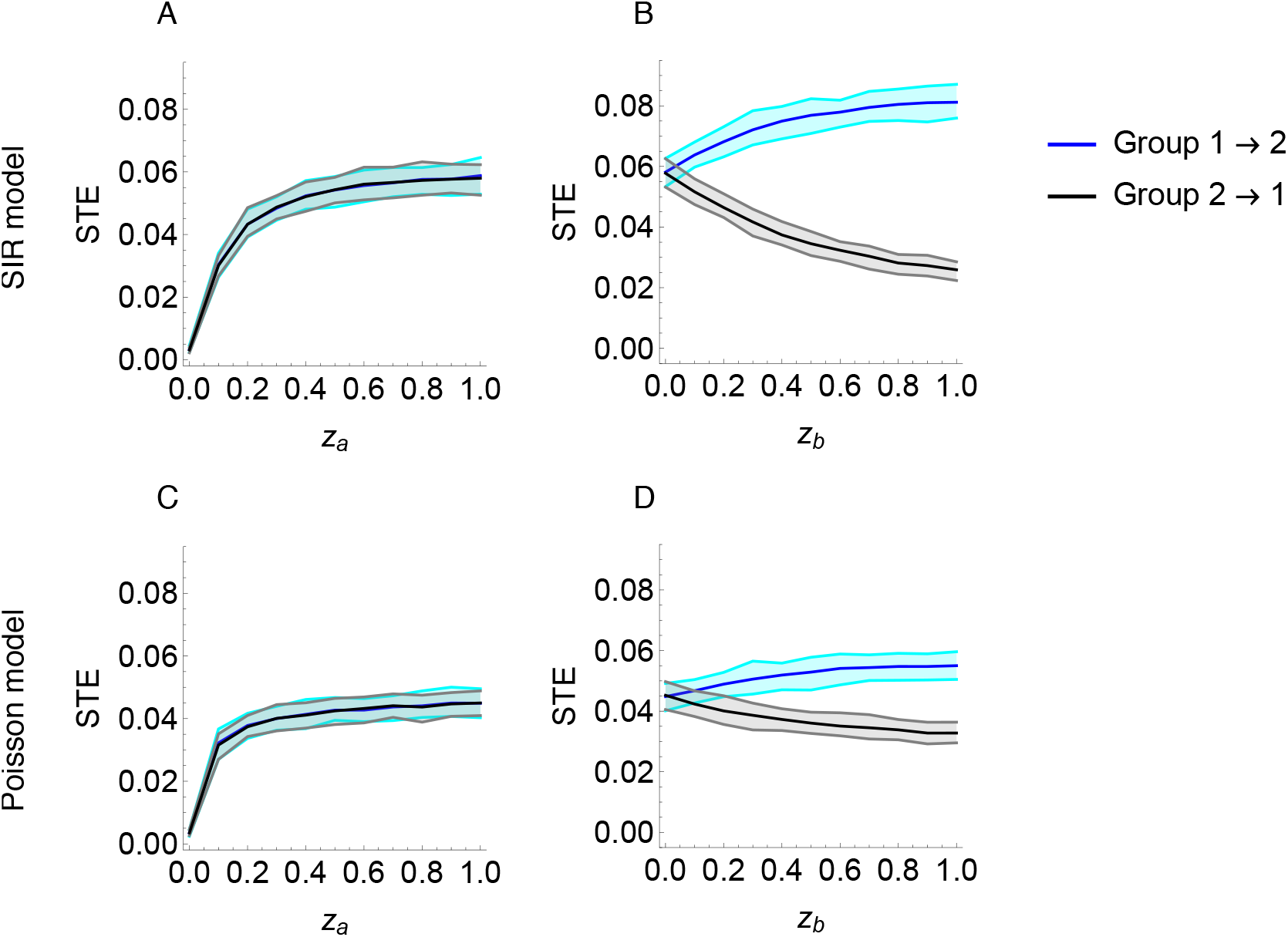
Mean (95% CI) Group 1 → 2 (blue) and Group 2 → 1 (black) STE values as the coupling between the two groups ranges from none to fully symmetric (A and C), and from fully symmetric to strongly driven by Group 1 (B and D). The curves are produced by simulating 100 ensembles of 800 epidemics each from the stochastic SIR model (A and B) or the Poisson model (C and D) for each value of *z*_*a*_ and *z*_*b*_ between 0 and 1 in steps of 0.1, and then calculating the between-group STE for each ensemble. The relative reproduction matrices that capture these two coupling scenarios are given in Eqs 3 and 4.

### 2.5. Reporting rates

Only a fraction of influenza cases are represented in the SDI-ILI dataset, since many people do not seek medical care for their symptoms. The tendency to seek medical care given infection with an ILI can vary by age group [43]. To factor this into the outbreak simulations, we introduce a reporting rate vector ***c*** in which element *c*_*i*_ gives the expected proportion of individuals in age class *i* who seek medical care when infected with an ILI. It is then possible to simulate a ‘reported’ disease incidence time series:

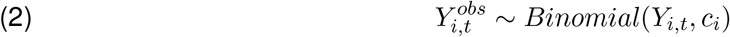

where *Y*_*i,t*_ is the simulated number of infected individuals in age class *i* at time *t* (under either model) and 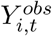 is the simulated reported number of infections in age class *i* at time *t*.

## 3. Results

### 3.1. STE reveals transmission asymmetries between two coupled age groups

We first calculate the STE between two age groups as the within- and between-group reproduction ratios vary. We consider between-group transmission that ranges from (a) fully decoupled to fully symmetric, and (b) fully symmetric to strongly driven by Group 1. The between-group infectiousness is specified using a “relative reproduction matrix” ***r***, which is a scaled version of the next-generation matrix [28], such that ***r***_*i,j*_*/****r***_*k,j*_ gives the proportional difference in group *j*’s infectiousness for group *i vs*. group *j*. For example, if ***r***_*i,j*_*/****r***_*k,j*_ = 2, then a member of group *j* is expected to infect twice as many members of group *i* than of group *k*. Scenario (a) is encapsulated by the relative reproduction matrix

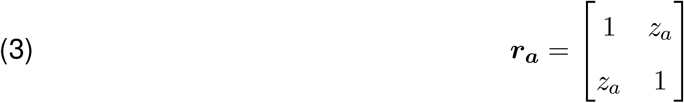

where *z*_*a*_ ∈ [0, 1]. Scenario (b) is encapsulated by the relative reproduction matrix

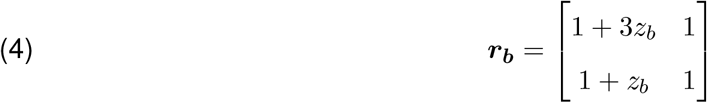

where *z*_*b*_ ∈ [0, 1].

Fig. 1 depicts the change in STE under these two transmission scenarios, calculated from epidemics simulated using the stochastic SIR model (Fig. 1 A–B) and the Poisson model (Fig. 1 C–D). Each pane in Fig. 1 is produced using 100 ensembles of 800 simulated epidemics for each value of *z*_*a*_ and *z*_*b*_ between 0 and 1 in steps of size 0.1. For each ensemble, the 800 simulated incidence time series are symbolised using symbols of length *m* = 3, and then the between-group transfer entropies are estimated using the relative symbol frequencies (see Fig. S3), producing 100 STE estimates for each value of *z*_*a*_ and *z*_*b*_. The solid blue (black) lines in Fig. 1 depict the mean Group 1→2 (Group 2→1) STE for each value of *z*_*a*_ and *z*_*b*_ across the 100 ensembles. The shaded blue (black) bands depict the range of the middle 95 Group 1→2 (Group 2→1) STE estimates for each value of *z*_*a*_ and *z*_*b*_ across the 100 ensembles, analogous to a 95% confidence interval. Under both the stochastic SIR and the Poisson models, the between-group STE increases steadily as the transmissive coupling ranges from none to symmetric (Fig. 1 A, C). Once Group 1 begins to dominate transmission, the Group 1→2 STE increases and the Group 2→1 STE decreases (Fig. 1 B, D), accurately capturing the transmissive relationship between the age groups.

When Group 1 drives transmission, the Poisson model yields a smaller difference in the STE between the two age groups than the stochastic SIR model does (Fig. 1 B, D). Visual inspection suggests that the simulated time series produced using the stochastic SIR model tend to feature more stochastic fluctuations than the time series produced using the Poisson model (Figs S7 and S8). Since STE is effectively a measure of how these stochastic fluctuations transmit from one age group to another, this may explain why the differences in STE calculated using the Poisson model are relatively less pronounced. Overall, the qualitative similarity between the STE estimates from the two transmission models suggests that the Poisson model is an acceptable approximation to the stochastic SIR model, and that simulations from the Poisson model tend to produce more conservative estimates of the difference in STE between age groups than the stochastic SIR model.

### 3.2. STE reveals transmission asymmetries despite incomplete reporting

Next, we evaluate how incomplete reporting influences the detection of asymmetries in transmission strength. Fig. 2 depicts the mean estimated STE across 100 ensembles of 800 epidemics each for reporting rates *c*_*i*_ between 0.1 and 1 in steps of 0.1, with equal reporting rates across all age groups. The epidemic simulations are produced using the Poisson model with relative reproduction matrix

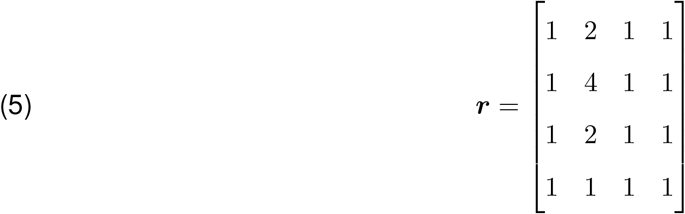

which could represent ‘children’ (Group 2) having strong within-group transmission (*r*_2,2_ = 4) and intermediate transmission to ‘infants’ (Group 1) and ‘adults’ (Group 3) (*r*_1,2_ = *r*_3,2_ = 2). Even for reporting rates as low as 0.1, the STE values from Group 2 are higher than those from any other group. As the reporting rates increase, the differences become more pronounced, accurately capturing the transmissive dominance of Group 2 over the other groups. The estimated STE increases with reporting rate for all age groups, but more quickly for Group 2 than for the other age groups. According to Biggerstaff *et al*. (2012) [43], true reporting rates for ILI in the US during the 2009 pandemic were between 0.4 and 0.6, for which the transmissive dominance of Group 2 is clear.

**Figure 2.**
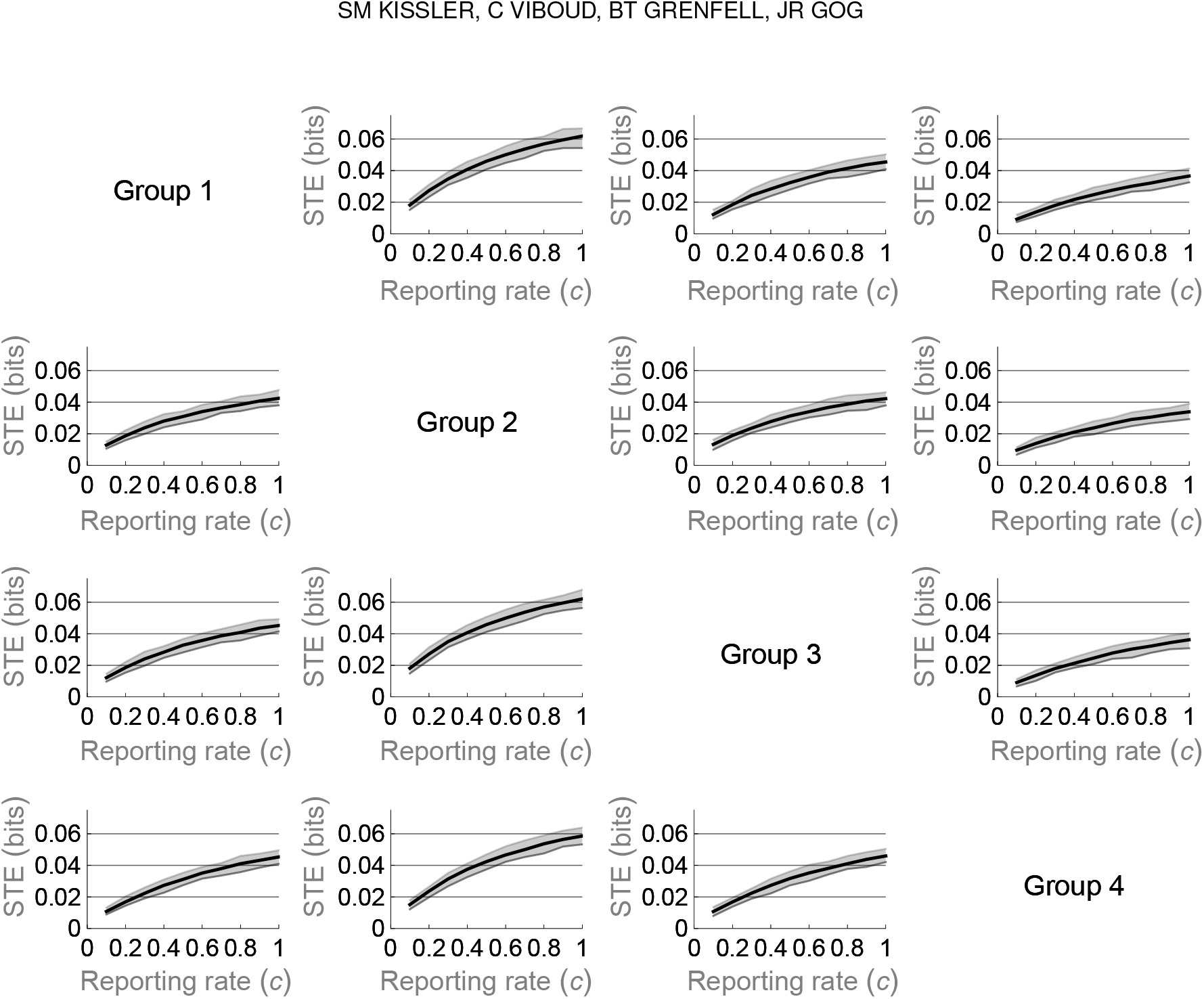
Mean pairwise STE values (solid lines) with 95% confidence intervals (shaded bands) for epidemics strongly driven by Group 2 under a range of reporting rates *c*. The curves are produced by simulating 100 ensembles of 800 epidemics each from the Poisson model for each value of *c* between 0.1 and 1 in steps of 0.1, and then calculating the between-group STE for each ensemble. The reporting rate *c*_*i*_ (see Eq. 2) is varied uniformly across all age groups *i*. The relative reproduction matrix that specifies within- and between-group transmission rates is given by Eq. 5. The plot in row *i* and column *j* depicts the STE from group *j* to group *i*.

### 3.3. STE reveals transmission asymmetries between twelve coupled age groups

To test the ability of STE to identify transmission asymmetries from data on the scale of the SDI-ILI dataset, we use the Poisson model to simulate 100 ensembles of 800 epidemics each with 12 age groups. We consider the scenarios (a) with the 12 × 12 relative reproduction matrix Eq. S48, representing high transmission from Groups 3–5 to Groups 3–5 (*r*_*i,j*_ = 4 for *i, j* ∈ {3, 4, 5}), intermediate transmission from groups 3–5 to groups 1–2 and 6–9 (*r*_*i,j*_ = 2 for *i* ∈ {1, 2, 6, 7, 8, 9} and *j* ∈ {3, 4, 5}), baseline transmission (*r*_*i,j*_ = 1) between all other groups, and uniform 50% reporting rate across all groups, and (b) with uniform transmission strength across all age groups (i.e. a 12 × 12 relative reproduction matrix with ‘1’ for all entries), 60% reporting rate for groups 1–5, and 40% reporting rate for groups 6–12, following the estimates of Biggerstaff *et al*. (2012) [43] for the ILI reporting rates in the United States during the 2009 influenza pandemic for children and adults, respectively.

Fig. 3 depicts the mean pairwise STE estimates between the 12 age groups under both scenarios. The square in row *i* and column *j* represents the STE from Group *j* to Group *i*. Darker squares correspond to higher STE. For the asymmetric transmission/uniform reporting rate scenario (scenario (a), Fig. 3A), the STE clearly captures the transmissive dominance of Groups 3, 4, and 5. The pairwise STE does not simply reproduce the structure of the relative reproduction matrix, as evidenced by the variability in mean pairwise STE for age groups other than Groups 3–5. This is because the STE captures a ‘knock-on’ effect for which information transferred from a strongly-driving age group can propagate through other age groups. For the uniform transmission/variable reporting rate scenario (scenario (b), Fig. 3B), it is evident that elevated reporting rates can also lead to elevated STE, both to and from the groups with elevated reporting rate (Groups 1–5). Overall, the variability in STE due to differences in reporting rate appears to be smaller than the variability in STE due to differences in transmission strength. Further discussion on the effect of reporting rates on STE may be found in the Supplemental Information.

**Figure 3.**
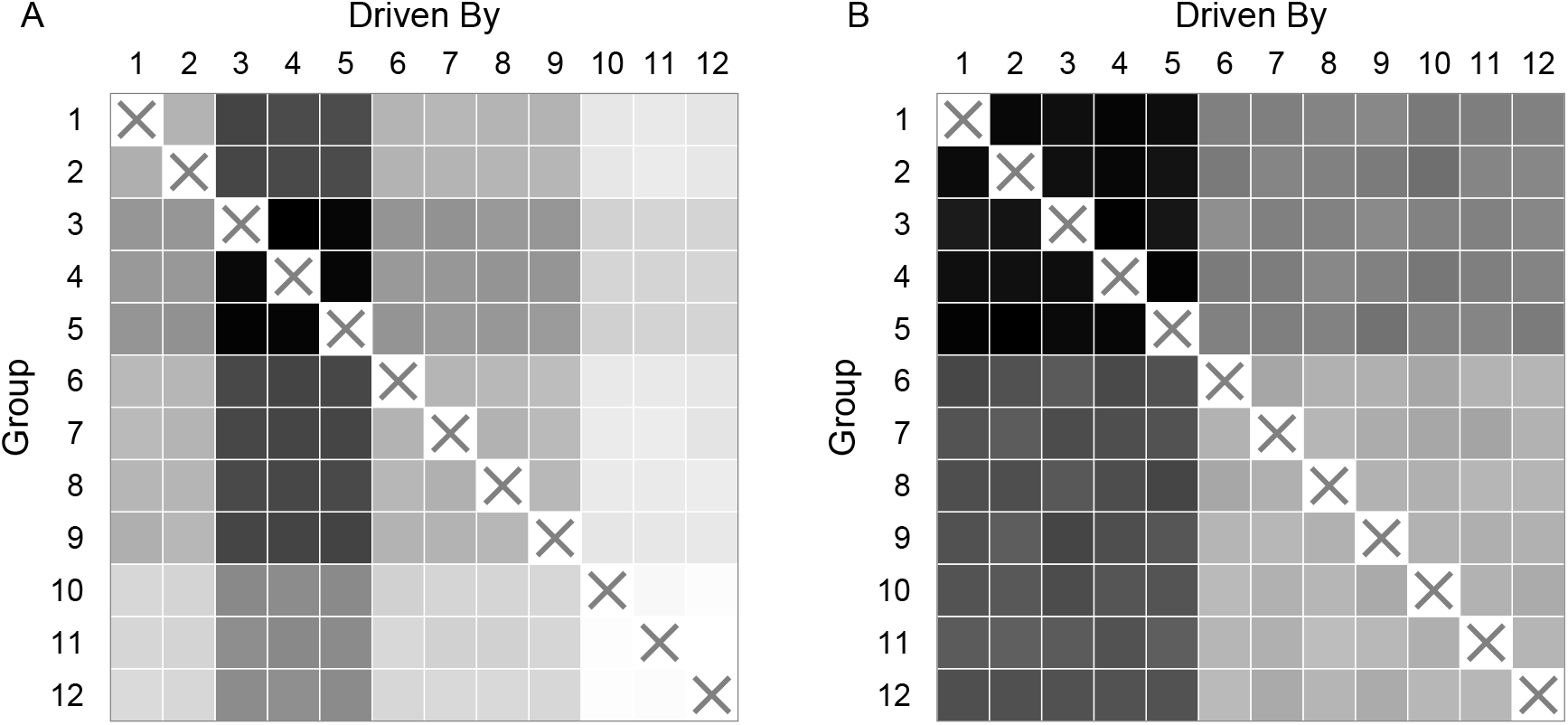
Mean pairwise STE values between 12 groups for epidemics strongly driven by Groups 3, 4, and 5 and uniform 50% reporting rate across all age groups (A), and for epidemics driven equally by all age groups, 60% reporting for Groups 1–5, and 40% reporting for Groups 6–12 (B). A box in row *i* and column *j* corresponds to the STE from group *j* to group *i*, where darker shades corresponds to higher STE. To generate the STE values, 100 ensembles of 800 epidemics were simulated from the Poisson model using relative rate matrix Eq. S48 for (A) or a relative rate matrix with all entries equal to 1 for (B). Each ensemble generates 144 pairwise STE values, so that each box represents the mean value across the 100 ensembles. The raw values are listed in Eqs S49 and S50.

### 3.4. School-aged children contributed disproportionately to transmission during the autumn 2009 A/H1N1pdm influenza outbreak in the US

To estimate the pairwise STE between the 12 age groups represented in the SDI-ILI dataset during the 2009 A/H1N1pdm influenza pandemic, we extract data from the 25 weeks between 12 July 2009 and 27 December 2009 and symbolise the ILI time series for each age group in each ZIP using a symbol length of *m* = 3. The pairwise STE values between all age groups are depicted in Fig. 4. The STE is highest in the columns representing 5–19 year-olds. This provides evidence that there was systematically elevated transmission from school-aged children to infants through adults. The adult-adult STE is also moderately elevated, suggesting that adults may have played a relatively important role in transmitting the outbreak amongst themselves, though this could also be explained by elevated transmission from children alone. Compare, for example, to the left-hand plot in Fig 3: in that simulation, only transmission from children is elevated, but it causes a moderate elevation in the STE from adults and infants to the other age groups due to the knock-on effect.

**Figure 4.**
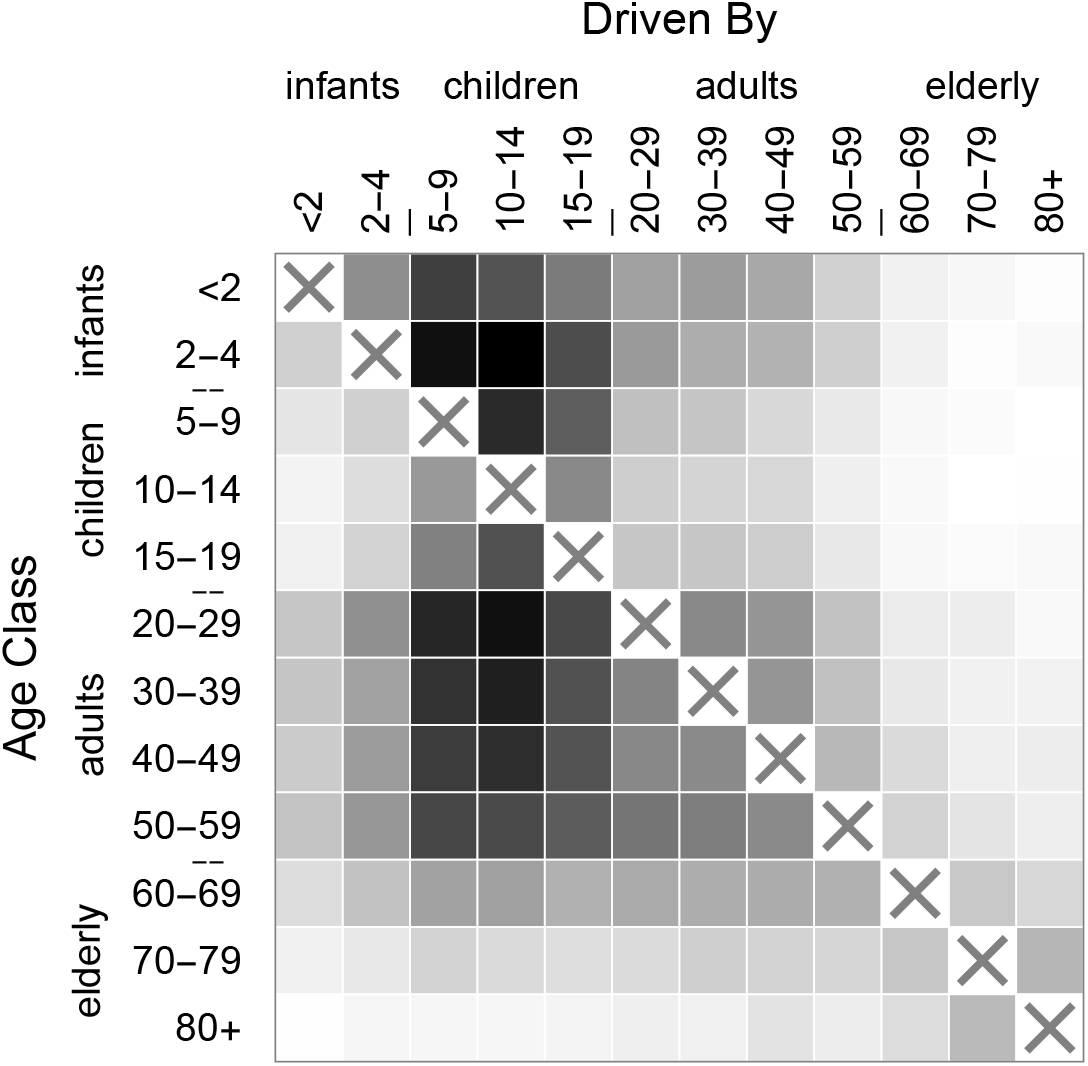
Mean pairwise STE values between the 12 groups represented in the SDI-ILI dataset during the autumn 2009 A/H1N1pdm pandemic influenza outbreak. A box in row *i* and column *j* corresponds to the STE from group *j* to group *i*, where darker shades corresponds to higher STE. The raw values are listed in Eq. S51.

As a control, we also calculated the pairwise STE between all age groups during 25 post-pandemic weeks, from 10 January 2010 through 27 June 2010. For these months, there is no apparent age structure in transmission (see Supplemental Information). We also calculated the pairwise STE between age groups for six previous influenza seasons (see Supplemental Information). For the 2009 pandemic, there is is a higher maximum pairwise STE and greater variation in the pairwise STEs than for any previous season. This could reflect differences in baseline ILI, which was likely lower during the autumn 2009 pandemic wave than during the seasonal outbreaks, due to the pandemic’s earlier timing. A lower baseline ILI might have made pairwise differences in STE easier to detect in 2009. However, the relatively higher and more heterogeneous STE values in 2009 are also consistent with the hypothesis that school-aged children played a disproportionately large role in the spread of the 2009 pandemic, as has been described elsewhere [4].

It is unlikely that differences reporting rates alone can account for the elevated STE from 5–19 year-olds to the other age groups. The mean pairwise STE values computed from simulations with uniform transmission rates and unequal reporting rates in Section 3.3 range from .0057 to 0.0084 (see Eq. S50), while the pairwise STE values computed from the SDI-ILI data range from 0.0056 to 0.084 (see Eq. S51), an order of magnitude larger. The mean pairwise STE values computed from simulations with asymmetric transmission and uniform reporting rates Section 3.3 range from 0.0047 to 0.014 (see Eq. S49), closer to the range observed from the SDI-ILI data but still somewhat smaller. This points towards a possible combined effect of strong transmissive driving from children plus elevated reporting in children. In addition, re-calculating the pairwise STE using probabilistic reconstructions of the pre-reporting SDI-ILI incidence time series (see Supplemental Information) indicate that the observed transmissive dominance of 5–19 year-olds persists even after adjusting for potential differences in reporting rate between children and adults. Furthermore, Biggerstaff *et al*. (2012) [43] report that 0-4 year-olds had the highest reporting rates for ILI in the United States in 2009, yet the STE from 0-4 year-olds is relatively low compared to the other age groups. If reporting rates alone could explain the observed differences in STE, the STE from infants should be at least as high as the STE from school-aged children.

It is also unlikely that the unequal partitions of the age groups can explain the observed patterns in the pairwise STE. The age groups under 20 years are partitioned such that they span fewer years, and thus contain fewer individuals, than the age groups above 20 years. Direct calculations and simulations (see Supplemental Information) indicate that, all else being equal, the out-going STE for a given group tends to increase as the group’s population size increases relative to the sizes of the other groups. If differences in the groups’ population sizes were driving the observed pairwise STE values, we would expect the age groups over 20 years to appear to dominate transmission – which is the opposite of what we observe here.

## 4. Discussion

Here, we propose STE as a means of ranking which age groups contribute most to the transmission of infectious disease outbreaks. STE is chosen for its robustness to point-wise noise and overall amplitude shifts in time series, which especially affect the ILI data stream due to non-influenza respiratory illness and incomplete reporting. Simulation studies indicate that STE can correctly rank transmissive asymmetries between age groups. However, STE is also positively associated with reporting rates, which can partially confound estimates of asymmetric transmission. STE estimates from ILI time series data from July-December 2009 in the United States suggest that 5–19 year-olds were primarily responsible for driving transmission of the autumn wave of the A/H1N1pdm pandemic influenza outbreak. It is unlikely that this result can be explained by differences in reporting rates alone.

The identification of school-aged children as the primary drivers of transmission of the 2009 influenza pandemic in the United States agrees with most other studies on age-specific transmission of both seasonal and pandemic influenza [4, 7, 8, 9]. Elevated transmission from school-aged children is likely due in part to the relatively high number of daily interpersonal contacts made by members of these age groups. Mossong *et al*. (2008) [8] for example estimate that 10–19 year-olds have more contacts per day than any other age group, and conclude from a modelling study based on empirical contact data that 5–19 year-olds are likely to both suffer the highest burden of disease and to drive the early-stage transmission of an outbreak transmitted by droplets through close contacts, like influenza. This underscores the importance of monitoring children during pandemic influenza outbreaks, and potentially prioritizing school-aged children for vaccination.

TE is closely linked to mutual information [31] and Granger causality [33]. Unlike TE, mutual information is symmetric; that is, it measures the probabilistic dependence between two processes, but cannot determine the direction of information transfer between them, if there is any [31]. Measuring the delayed mutual information between two processes is one way to introduce asymmetry. This takes a step toward inferring whether one process influences another, by measuring shared information between the present state of one process and the past states of another [31]. While the lagged mutual information describes how one process’ history predicts the static probabilities of another, the TE measures how one process’ history influences the transition probabilities of another. Because of this, the TE is less likely to be confounded by a shared input signal, and is a better measure of stochastic ‘driving’ [31]. Section 2 of Kaiser and Schreiber (2002) [44] provides a detailed description of the differences between TE and mutual information. Granger causality, on the other hand, is a special case of TE that arises when the stochastic processes are jointly Gaussian-distributed [45]. The TE is thus better suited than Granger causality for making inferences on more general, possibly nonlinear, processes, though this comes at the expense of requiring more data and having no clear way to test statistical significance [45].

Convergent cross mapping (CCM) [34] was developed to solve a similar problem as TE, but is based on somewhat different underlying theory. CCM was developed to detect so-called ‘causal’ relationships in partially stochastic systems with underlying deterministic structure. CCM relies on Takens’ theorem [46] to reconstruct candidate manifolds of the underlying dynamical system using lagged observations from two time series. ‘Causality’ is inferred if nearby points on one reconstructed manifold consistently map to nearby points on the other reconstructed manifold. CCM has been used to provide evidence that temperature and absolute humidity fluctuations drive the timing of global seasonal influenza outbreaks [47], though some controversy surrounds these findings [48, 49]. Nevertheless, it would be interesting to see whether CCM can reveal asymmetric epidemiological interactions between age groups, and to compare its findings with those identified using TE. Lungarella *et al*. (2007) [50] provide more detail on the relationships between various methods that infer asymmetric relationships from time series data. (As an aside, we prefer to avoid the term ‘causality’ with respect to these methods, despite its frequent use in the literature. Regardless of the vocabulary used, they have successfully detected meaningful relationships between real-world coupled dynamic processes [30, 32, 34, 51, 52, 53]).

Despite the apparent well-suitedness of STE for making inferences from ILI data, its epidemiological relevance currently remains limited. The calculation of STE requires no prior epidemiological information whatsoever, which makes its success somewhat surprising. The next-generation matrix [28] is the key object for characterising age-structured, or more generally population-structured, disease transmission dynamics, and yet there is no obvious direct link between STE estimates and the NGM. It is possible that further simulation studies could help identify such a link; even though the STE values seem to bear little mechanistic meaning apart from the relative ordering of age groups that they yield, it is possible that regressing the inferred STE values on an underlying known NGM could connect the pairwise STE matrix with the NGM under certain conditions. However, it appears unlikely that a simple link exists, especially since STE can say nothing about transmission within a single age group, which is necessary for filling in the diagonal entries of the NGM. STE and related methods such as CCM that do not explicitly incorporate mechanistic descriptions of the underlying physical system are unlikely to be able to reveal more than an approximate hierarchy of driving processes. Nevertheless, such a hierarchy can contain valuable information, especially if developing and fitting a mechanistic model is too demanding to be practicable. Certain extensions to STE could also enhance its relevance for epidemiological inference. Local transfer entropy [54] and state-dependent transfer entropy [55], like the contextual STE (see Supplemental Information), are intended to make the TE more flexible and general, by considering how information transfer may change under varying conditions or ‘meta-states’. These extensions may yield better insight into epidemic processes, which are inherently nonlinear and context-dependent, than the more traditional measurements of transfer entropy can provide.

Perhaps the most important challenge confronting the TE and related measurements is deciding how to measure statistical power and significance. STE calculations rely on a middle level of stochasticity in the underlying stochastic processes; for a deterministic system, the STE will always be exactly zero, while for a stochastic system with too much within-sequence noise, the small-scale variation in amplitudes will likely mask important patterns from which the transfer of information might be inferred. The acceptable range of stochasticity has not been clearly defined. Similarly, it is unclear how best to measure when a difference in STE should be called statistically significant. Though this is recognised as an open and difficult problem [45, 48], it may be possible to make some progress by assuming that the underlying process follows certain epidemiological, or otherwise well-specified, dynamics.

## 5. Disclaimer

This paper does not necessarily represent the views of the US government or the NIH.

## Data Availability

Data will be available from https://github.com/skissler/STE upon publication

